# The Construction of Multi-ethnic Polygenic Risk Score using Transfer Learning

**DOI:** 10.1101/2022.03.08.22272114

**Authors:** Zhangchen Zhao, Lars G. Fritsche, Jennifer A. Smith, Bhramar Mukherjee, Seunggeun Lee

## Abstract

As most existing genome-wide association studies (GWAS) were conducted in European ancestry cohorts and as the existing PRS models have limited transferability across ancestry groups, PRS research on non-European ancestry groups is negatively impacted. Here we propose a novel PRS method using transfer learning techniques. Our approach, TL-PRS, uses gradient descent to fine-tune the baseline PRS model from an ancestry group with large sample GWAS to the dataset of target ancestry. In our application of constructing PRS for the six quantitative and two dichotomous traits for 10,285 South Asian and 8,168 African ancestry individuals in UK Biobank, TL-PRS achieved up to 42% average relative improvement compared to the existing methods. Our approach increases the transferability of PRSs across ancestries and thereby helps reduce existing inequities in genetics research.

## INTRODUCTION

Genetic risk prediction is one of the widely investigated topics in genetic epidemiology as it can help us better understand the genetic architecture of complex traits and potentially aid clinical decision-making^1-3^. Many polygenic risk score (PRS) construction methods have been developed, including pruning and thresholding (PT)^4^, Lassosum (Lsum)^5^, PRS-CS^6^ and LDpred^4^. Overall, these methods perform well and help to identify high risk groups within the same ancestry group^2,4,7,8^. However, due to insufficient GWAS data from non-European ancestry groups such as South Asian and African ancestry, PRSs for these diverse ancestry groups often only show limited prediction performance^5,6^. In addition, due to genetic differences across ancestry groups, the direct use of PRS models trained with European data to non-European individuals was shown to lead to reduced prediction accuracy ^4,8^.

To address this issue, Márquez□Luna et al. proposed a multi-ethnic PRS model by linearly combining two PRSs, each trained from different ancestry GWAS summary statistics^9^. They attained more than 70% relative improvement in prediction accuracy for type 2 diabetes in both Latino and South Asian ancestry cohorts compared to prediction models from a single ancestry GWAS. PRS-CSx^10^ implemented the same linear combination approach using two PRSs trained with PRS-CS. However, this linear combination approach implicitly assumes that the optimal effect sizes (or beta coefficients) weighting for prediction is a linear combination of the effect sizes of two PRSs, which may not hold in all situations. In addition, this method cannot be used when GWAS summary statistics are only available for one ancestry.

Here we propose a novel multi-ethnic PRS using transfer learning techniques^11^, borrowed from the machine learning literature. Transfer learning is a widely used tool that applies an existing trained model to a different but related problem. The usual procedure of transfer learning is a gradient-based optimization when modeling the second task.^12,13^ From the practical viewpoint, the reuse or transfer of information from previously learned tasks for the learning of new tasks has the potential to significantly improve the prediction performance compared to the baseline methods as well as reduce the required sample size of training data^11^.

Our approach, Transfer Learning PRS (TL-PRS), fine-tunes the baseline model trained with GWAS summary statistics from a larger sample size ancestry group to a smaller target ancestry group. TL-PRS can use PRSs from any existing PRS methods (such as Lsum and PRS-CS) as a baseline model. Using the effect sizes of the baseline model as initial values, TL-PRS iterates the gradient descent algorithm to adapt the effect sizes for the target ancestry group. In the presence of multiple GWAS summary statistics from different ancestries, TL-PRS fine-tuned linearly combined PRS. Since TL-PRS uses a simple gradient descent, it is scalable for large cohort datasets.

In our simulations, TL-PRS outperformed existing PRS methods in a wide range of genetic architectures and cross-ancestry genetic correlations. In a real-world example with individual level data of the UK-Biobank (UKBB), we use an European ancestry GWAS from UKBB and East Asian ancestry GWAS from Biobank Japan (BBJ) as training data to predict eight traits in 10,285 South Asian and 8,168 African ancestry samples. Compared to the baseline methods, TL-PRS substantially improved the prediction accuracy for most traits. For example, TL-PRS obtained an 13-42% average relative improvement for African samples compared to the baseline methods. By improving the polygenic risk prediction in non-European ancestry individuals, our approach will help reduce the prevailing inequities in genetic and health research.

## RESULTS

### Overview of TL-PRS

We first build PRS models using existing methods, and these models provide effect size estimates of genetic variants, which are used as initial values of TL-PRS. In this paper, we used Lsum^5^ and PRS-CS^6^ trained models as the baseline methods, which are referred as TL-PRS-Lsum and TL-PRS-CS, respectively. TL-PRS method can also be applied to any other training models, such as LDpred^4^. When more than one summary source is available, we can linearly combine the baseline models first as the initial value and then implement transfer learning (referred as MTL-PRS).

The hyperparameters in TL-PRS include the learning rate and the number of iterations. Given TL-PRS models from different GWAS summary sources, we can integrate them by learning an optimal linear combination and then use it as the initial value to implement TL-PRS (Figure 1). Figure 2 shows the relative accuracy 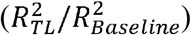 of TL-PRS as a function of iterations. The relative accuracy in the training dataset continue to increase as the number of iterations increases, which caused the overfitting. However, the fifth iteration reached the maximum relative accuracy in the validation sets of both simulation and real data analysis, which suggested that the fifth iteration was the optimal point to stop in these two examples. A similar strategy can be applied to choose the learning rate.

**Figure 1:**
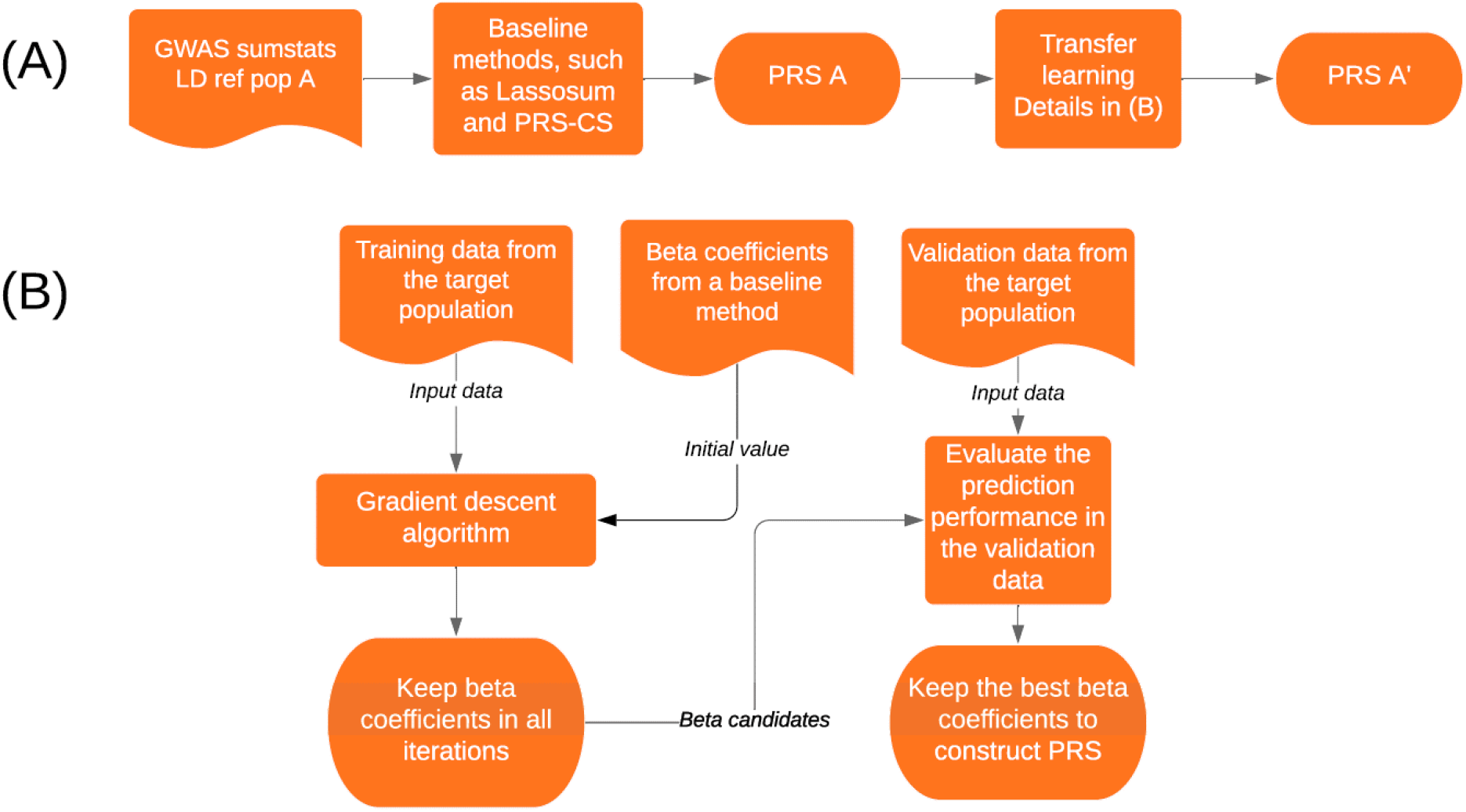
Overview of TL-PRS methods. LD ref: LD reference panel. (A) The general procedure to construct TL-PRS; (B) The detailed procedure of TL-PRS. The training data from the target population does not need to be individual-level data. Validation data is recommended to be individual-level data to achieve the best prediction performance.

**Figure 2.**
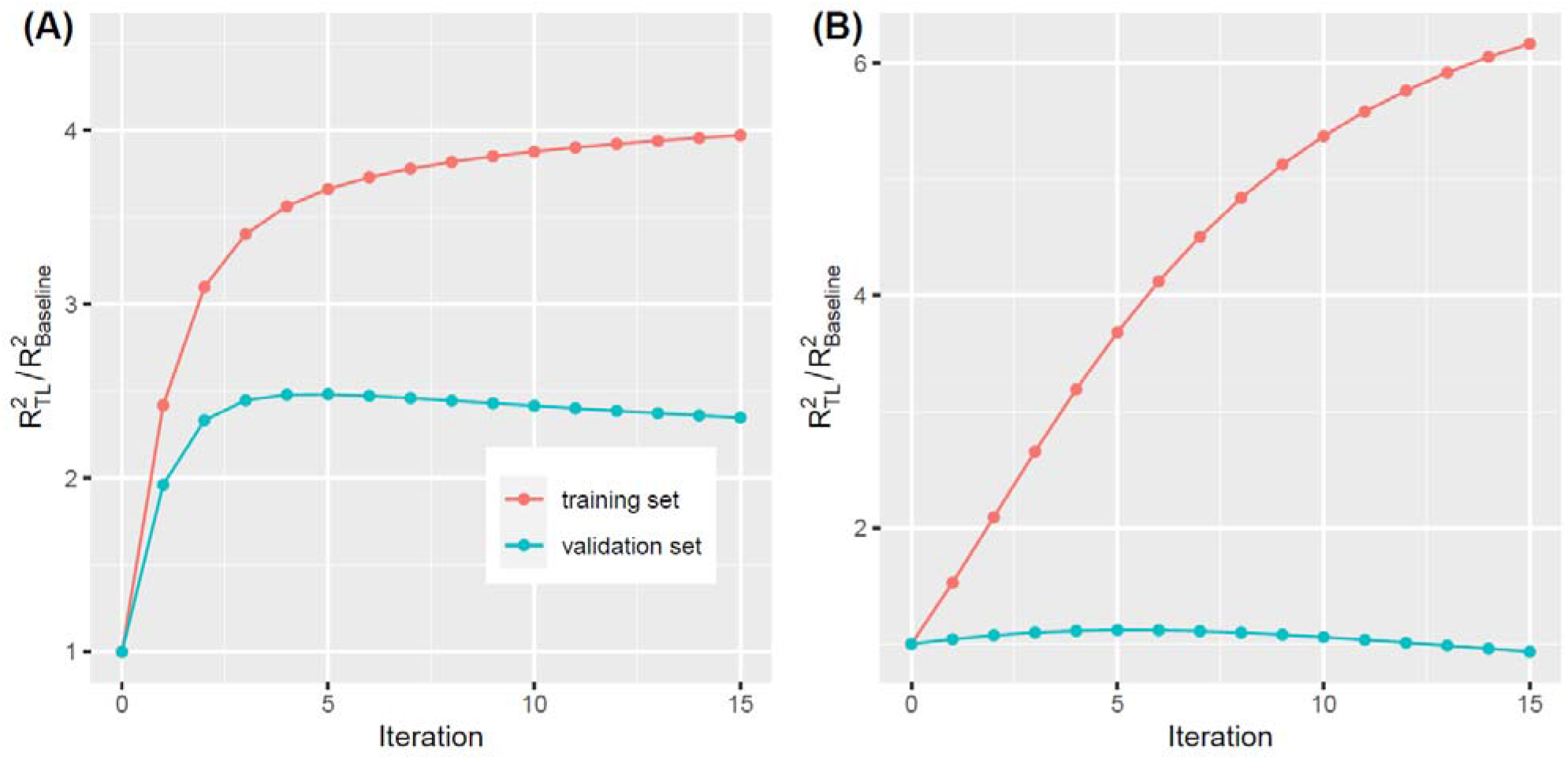
Relative accuracy of transfer learning method by the number of iterations. (A) The simulation setting where the causal markers were 0.1%, genetic correlation was 0.4 and European summary statistics were used. (B) The real data analysis of HDL in a South Asian cohort from UK Biobank, where UKBB summary statistics were used.

### Simulations using South Asian samples in the UK Biobank

In the simulation, different scenarios were considered by randomly selecting 0.1% or 1% variants across the genome as causal variants, which explained 50% of the phenotypic variance in total. Additionally, causal variants were assumed to be the same across ancestry groups, but different effect sizes were simulated from a multivariate normal distribution using the cross-ancestry genetic correlation 0.4, 0.7 and 1^10^. We generated 20 datasets in each scenario to evaluate the predictive performance of different PRS construction methods. We evaluated single-source prediction methods (PT, Lsum, TL-PRS-Lsum, PRS-CS, and TL-PRS-CS) that use a single ancestry group GWAS to build prediction models and multi-source prediction methods (PT-multi, Lsum-multi, MTL-PRS-Lsum, PRS-CSx, and MTL-PRS-CS) that utilize multiple ancestry group GWAS. The implementation details can be found in Table S1.

#### Results of single-source polygenic prediction methods in simulation

The prediction accuracy of single-source and multi-source polygenic prediction methods in the simulations can be found in Figure 3. For a fixed heritability 0.5, the predictive performance of all ten PRS methods decreased when the genetic architecture became more polygenic (0.1% vs 1% causal). Although the causal variants were identical across the ancestries, all ten PRS methods showed decreased prediction accuracy when the genetic effects were less correlated among ancestries. This is also the situation where TL-PRS could further improve the prediction accuracy. For example, when genetic correlation was 0.4, TL-PRS-Lsum improved a 241% and 57.1% average prediction accuracy compared to Lsum when the causal variants were 0.1% and 1%, respectively (Figure 4). The relative improvement of TL-PRS-CS over PRS-CS was 44.4% and 44.8% on average. However, when genetic correlation was 1.0, Lsum and PRS-CS are sufficient for prediction in target ancestry because the training and testing data shared same true effect sizes. TL-PRS-Lsum and TL-PRS-CS could attain limited relative improvement of the prediction accuracy in this situation. In general, TL-PRS performed better when the genetic correlation was smaller and when the causal variants were sparser.

**Figure 3.**
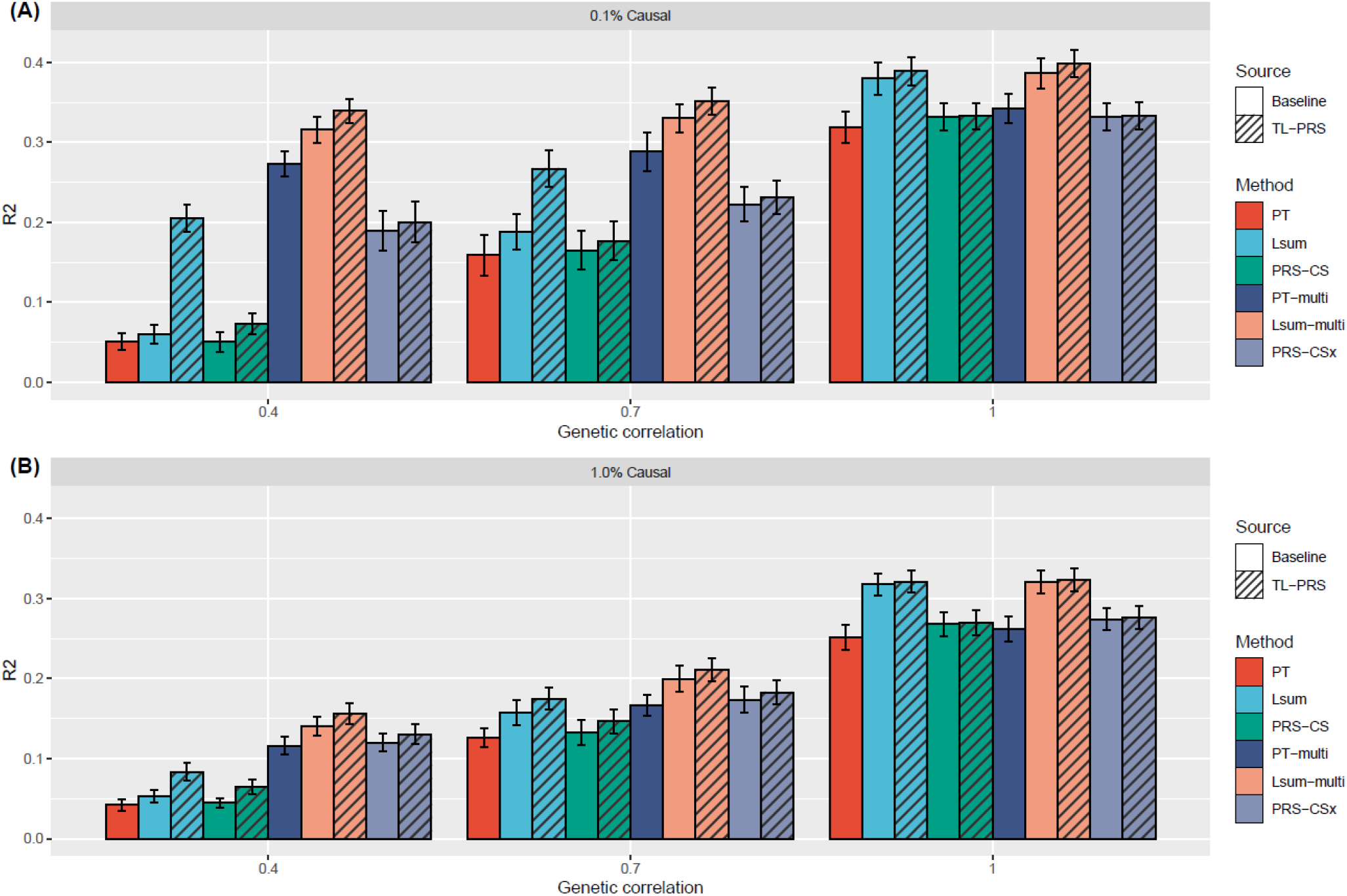
Prediction accuracy of single-source and multi-source polygenic prediction methods in simulations. Two different percentages of causal variants (0.1% and 1% causal variants) and three different cross-population genetic correlations (0.4, 0.7 and 1.0) were considered. Heritability was fixed at 50%. Prediction accuracy was measured by the squared correlation (R2) between the simulated and predicted phenotypes in the testing dataset, averaged across 20 simulation replicates. Error bar indicates the standard deviation of R2 across simulation replicates.

**Figure 4.**
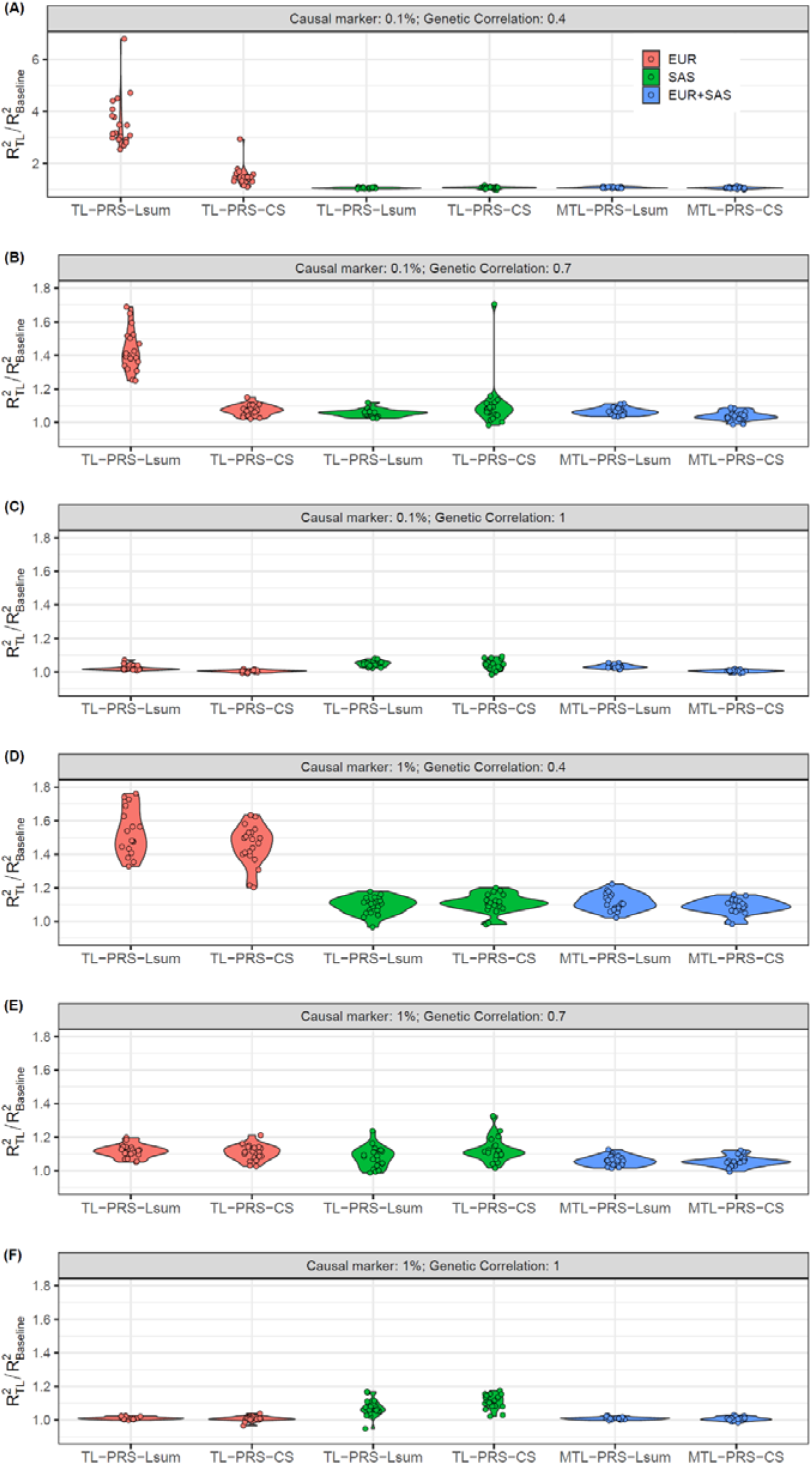
Relative prediction accuracy of single-source and multi-source TL-PRS, with respect to the baseline models across 20 replicates in the simulation. Note the maximum value of Y-axis is 7 for (A) and 1.8 for all other plots.

#### Results of multi-source polygenic prediction methods in simulation

We further assessed whether multi-source prediction methods (PT-multi, Lsum-multi, MTL-PRS-Lsum, PRS-CSx, MTL-PRS-CS) could improve cross-ancestry polygenic prediction. Specifically, we combined PRS models from European ancestry summary statistics (N=100K) and South Asian ancestry summary statistics (N=10K). When the genetic correlation was 1, the multi-source prediction methods cannot improve prediction accuracy in comparison with the single-source prediction methods using European ancestry summary statistics, because European ancestry shared the same true effect sizes as South Asian ancestry and had ten times sample size. In the scenario where the genetic correlation was less than 1, multi-source prediction methods improved prediction accuracy over single-source prediction methods, reflecting the increase in source sample size. Overall, while Lsum-multi outperformed PT-multi and PRS-CSx in most cases, MTL-PRS-Lsum further improved cross-ancestry prediction accuracy comparing Lsum-multi across all simulation settings (Figure 3 & 4). For example, when genetic correlation was 0.4, TL-PRS-Lsum improved a 7.38% and 11.6% average prediction accuracy compared to Lsum-multi when the causal variants were 0.1% and 1%.

In the training step, TL-PRS does not require individual-level data as the gradients can be calculated with summary statistics. To evaluate whether using summary statistics can reduce the performance of TL-PRS, we compared it with a TL-PRS version with individual-level data, TL-PRS(ind). Table S2 compares the model requirements of TL-PRS and TL-PRS(ind). Figure S1 further showed that TL-PRS had similar prediction R^2^ compared to TL-PRS(ind), which shows summary statistics are sufficient for TL-PRS training.

Overall, our simulation shows that TL-PRS-Lsum and TL-PRS-CS robustly improves cross-ancestry prediction over PT, Lsum and PRS-CS across varying genetic architectures and genetic correlations. The relative improvement of TL-PRS compared to the baseline method is over 40% when genetic correlation is 0.4.

### Prediction performance for South Asian and African ancestry samples in the UK Biobank

After excluding related individuals, the target sample size of South Asians (SAS) and Africans (AFR) were 10,285 and 8,168, respectively. We randomly split them into training dataset (for model fitting), validation dataset (for hyper-parameter tuning) and testing dataset (for the evaluation of predictive performance) (Table S3). We applied single-source prediction methods to the UKBB or BBJ GWAS summary results and used multi-source prediction methods to combine the UKBB and BBJ GWAS results.

Table 1 shows the prediction accuracy of different PRS construction methods in analyses of LDL in the African cohort of UK Biobank. When using UKBB GWAS results, the prediction R^2^ of TL-PRS-Lsum (0.058) and TL-PRS-CS (0.028) were higher than Lsum (0.033) and PRS-CS (0.022). In addition, when using BBJ GWAS results, the prediction R^2^ of TL-PRS-Lsum (0.068) and TL-PRS-CS (0.028) was higher than Lsum (0.048) and PRS-CS (0.023), demonstrating higher prediction accuracy in TL-PRS models. When combining UKBB and BBJ GWAS results, both Lsum-multi (0.052) and PRS-CSx (0.037) outperformed PT-multi (0.030), as expected. At the same time, MTL-PRS-Lsum (0.068) and MTL-PRS-CS (0.044) reached the best prediction accuracy. The consistent conclusions were reached when using the criteria of beta coefficients of normalized PRS or the mean difference between top 10% and bottom 10% PRS. The detailed results of other traits in the SAS and AFR ancestries can be found in Table S4 and S5, respectively.

**Table 1.** Prediction accuracy of 15 different approaches to construct PRS in the analyses of LDL in the African cohort of UK Biobank. For single source PRS methods, the training GWAS summary source is shown in the parentheses. Best prediction R2 is marked as bold.

| Model | Prediction R2 of PRS | Mean difference between top 10% and bottom 10% PRS |
| --- | --- | --- |
| **Single-source PRS methods** |  |  |
| PT (UKBB) | 0.012 | 0.388 |
| Lsum (UKBB) | 0.033 | 0.625 |
| TL-PRS-Lsum (UKBB) | 0.058 | 0.779 |
| PRS-CS (UKBB) | 0.022 | 0.552 |
| TL-PRS-CS (UKBB) | 0.028 | 0.506 |
| PT (BBJ) | 0.028 | 0.474 |
| Lsum (BBJ) | 0.048 | 0.595 |
| TL-Lsum (BBJ) | 0.068 | 0.795 |
| PRS-CS (BBJ) | 0.023 | 0.421 |
| TL-PRS-CS (BBJ) | 0.028 | 0.565 |
| **Multi-source PRS methods** |  |  |
| PT-multi | 0.030 | 0.531 |
| Lsum-multi | 0.052 | 0.727 |
| MTL-PRS-Lsum | **0.068** | **0.926** |
| PRSCS<sub>x</sub> | 0.037 | 0.662 |
| MTL-PRS-CS | 0.044 | 0.721 |

Consistent with the simulation results, TL-PRS-Lsum and TL-PRS-CS outperformed Lsum and PRS-CS in most traits from SAS and AFR (Figure 5). For SAS ancestry, TL-PRS-Lsum attained 8% and 2% average relative improvement in prediction accuracy using BBJ and UKBB GWAS results compared to Lsum; the relative improvement of TL-PRS-CS over PRS-CS was on average 39% and 10% respectively. For AFR ancestry, TL-PRS-Lsum attained 13% and 34% relative improvement of prediction accuracy in BBJ and UKBB GWAS results compared to Lsum; TL-PRS-CS improved prediction accuracy by 42% and 23% compared to PRS-CS. When combining BBJ and UKBB GWAS results, MTL-PRS-Lsum and MTL-PRS-CS had higher prediction performance than Lsum-multi and PRS-CSx (Figure 5).

**Figure 5.**
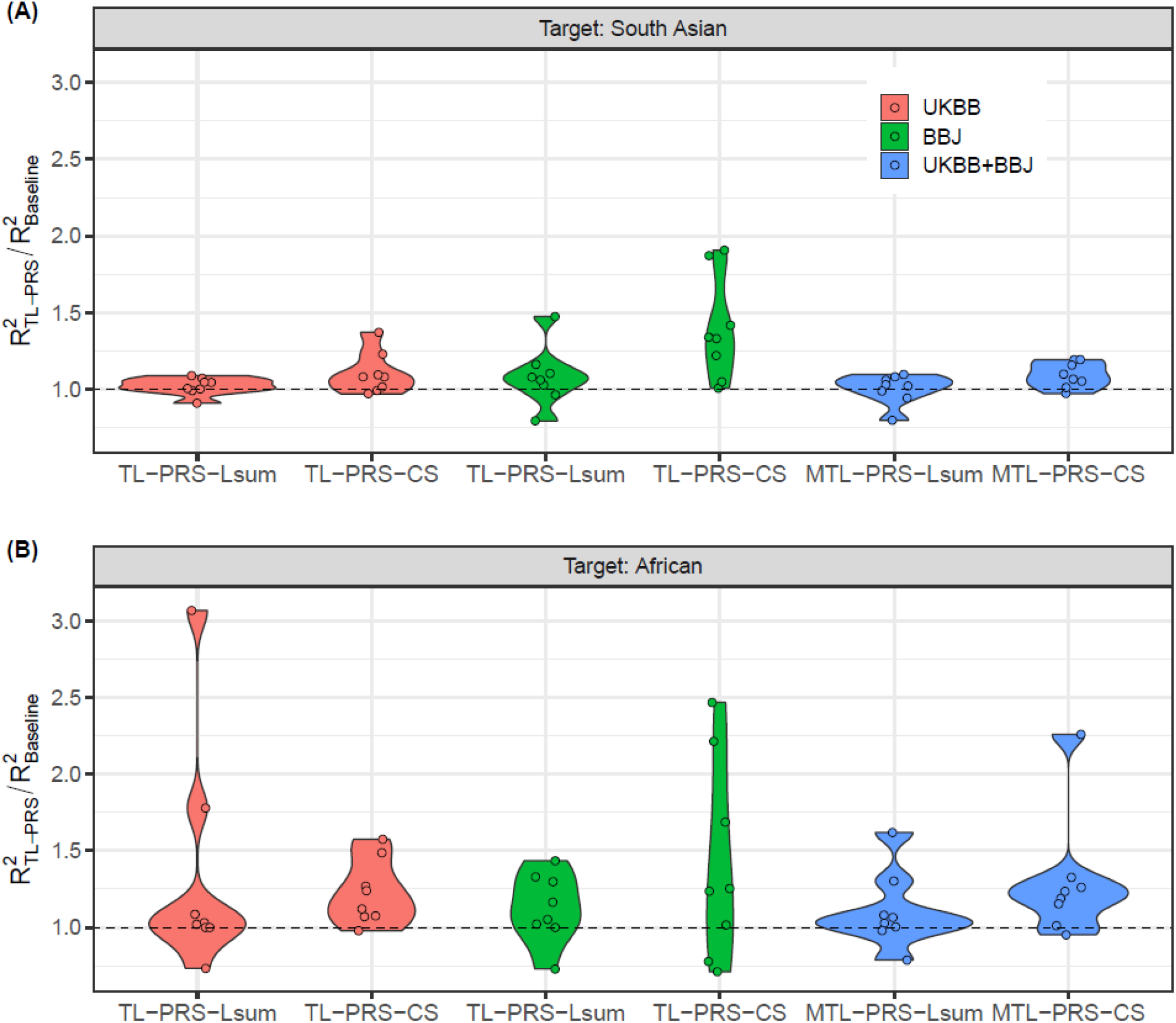
Relative prediction accuracy of single-source and multi-source TL-PRS, with respect to the base models without transfer learning across 8 traits in South Asian and African. Each point shows the relative prediction R^2^ of a trait.

Figure 6 further compares all ten PRS methods among all eight traits in the South Asian and African ancestry individuals. This bar plot summarizes the number of times each PRS method ranked top 3 in terms of prediction R^2^ for 16 traits and ancestry combinations (8 traits × 2 ancestries). (The detailed comparison can be found in Table S6.) Compared to the baseline methods, TL-PRS always appeared more times in the top 3 than the baseline method, demonstrating the ability of TL-PRS to improve prediction accuracy. In addition, MTL-PRS generally performed better than TL-PRS because MTL-PRS incorporated two different ancestries. Overall, MTL-PRS-CS shows the most robust performance across all situations since it ranks top 3 in almost all situations (15/16).

**Figure 6.**
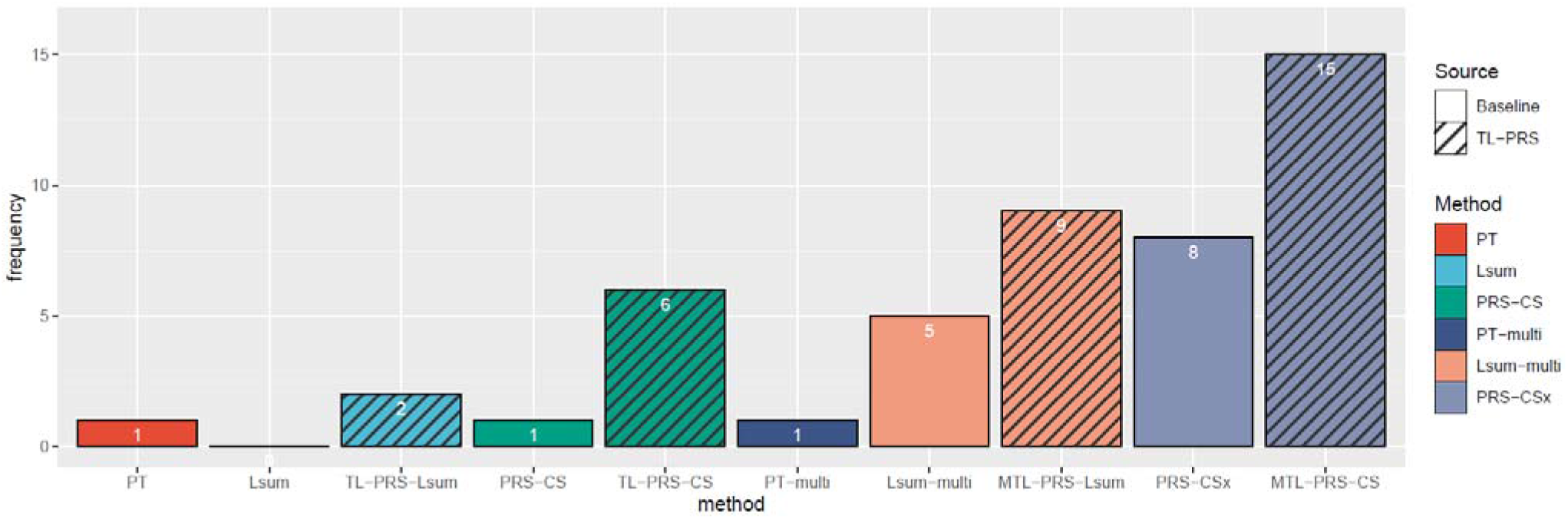
The method comparison for all eight traits in the South Asian and African ancestry individuals. Y-axis represents the number of times that each PRS method ranked top 3 in terms of prediction R^2^ for 16 traits and ancestry combinations (8 traits × 2 ancestries). Single-source prediction methods (PT, Lsum, TL-PRS-Lsum, PRS-CS, TL-PRS-CS) based on UKBB and BBJ GWAS results and multi-source PRS methods (PT-multi, Lsum-multi, MTL-PRS-Lsum, PRS-CSx, MTL-PRS-CS) were included in the comparison.

Figure S2 shows the cumulant event plot using the samples in the top 10% PRS across two ancestries. Across all situations, TL-PRS methods were found to have a similar or higher cumulant event curve than the baseline method. For example, in the analysis of CAD in the AFR cohort, when the age was up to 70, the cumulant prevalence of the samples with the top 10% PRS constructed by TL-PRS-Lsum(UKBB) was 0.16 while the prevalence in the samples with the top 10% PRS using Lsum(UKBB) was 0.12, suggesting that the TL method can improve the prediction of individualized disease risk and trajectories.

In general, compared to the baseline methods, TL-PRS obtained 2-39% average relative improvement for SAS ancestry individuals; 13-42% for AFR ancestry individuals. Among all ten PRS methods, MTL-PRS-CS is recommended due to its robust performance across all possible situations.

## DISCUSSION

We have presented the TL-PRS method, which can adapt the PRS model from other ancestries to the target ancestry. We have shown, through simulation studies, that TL-PRS-Lsum and TL-PRS-CS robustly improves cross-ancestry prediction over Lsum and PRS-CS across traits with varying genetic architectures, genetic correlations between target ancestry and samples used for calculating summary statistics. Using both quantitative and dichotomous traits from SAS and AFR ancestries in UK Biobank, we have demonstrated the TL-PRS can leverage large-scale European ancestry GWAS to boost the accuracy of polygenic prediction in non-European ancestries, for which ancestry-matched GWAS results may be orders of magnitude smaller in sample size.

Overall, the performance of TL-PRS depends on many factors, such as target ancestries, trait types, and baseline methods. When genetic correlations between target ancestry and samples used for calculating summary statistics are large, the baseline methods are sufficient for prediction and TL-PRS might not further improve the prediction performance. When genetic correlations are small, TL-PRS can be applied on the target data to help adapt effect sizes of the existing model to the target data. TL-PRS appears to be most effective when genetic correlations are small. MTL-PRS-CS is recommended in general due to its robust performance in real-data analysis. Moreover, the approach could be further extended to admixed populations with simple modifications. Future work is needed to better evaluate the performance in admixed populations.

TL-PRS can use GWAS summary results of the target samples to calculate gradients for transfer learning. In our simulation and real-data analysis, we only used GWAS summary results for TL-PRS training. However, TL-PRS still requires individual level data for validation and testing datasets. When the individual level data of validation dataset is not available, pseudo-validation^5^ could be applied for tuning the hyperparameters but the performance is unstable^14^.

Despite these advantages, our work is subject to limitations and leaves several questions open for future exploration. First, although we have demonstrated large relative improvements in prediction accuracy, absolute prediction accuracies are not sufficiently high to achieve clinical utility for most traits ^15,16^; our simulations suggest that multi-ethnic polygenic risk scores will continue to produce improvements when more diverse GWAS results are available, and the sample sizes are larger. Second, when combing two summary sources, the improvement of our MTL-PRS over the existing best PRS methods (PT-multi, Lsum-multi and PRS-CSx) is limited. More research work is needed to combine more than one summary source. For example, the heritability of the traits, which may differ across ancestries due to environmental factors, such as health behaviors and socioeconomic factors, can also be used to tune the weights for their linear combination. Additionally, we did not incorporate data from the X chromosome, which is likely to harbor additional heritability that could improve prediction in some traits^17^. Finally, we restricted our analyses on common variants, but we may wish to incorporate the effects of rare variants in the future work.

While extending present research to acquire more diverse ancestry genomes with sample sizes equivalent to European ancestry samples is the optimal, in the meantime, all existing available information should be efficiently used to improve prediction across ancestries. We believe that TL-PRS can increase the usefulness of PRS in multiple ethnic groups and reduce potential health inequities.

## METHODS

### Polygenic risk score construction using single ancestry GWAS summary statistics

With GWAS summary statistics (i.e. the effect size estimate and standard error), a PRS is constructed as the summation of the estimated effects across all genetic variants on a given phenotype. For individual *i*, PRS can be defined as

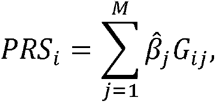

where *M* is the number of variants, *G*_*ij*_ is the genotype of the genetic variant *j*, and 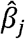 is the effect size. There are several well-known methods that estimate the effect sizes 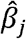 using GWAS summary and linkage disequilibrium (LD) information, such as pruning and thresholding (PT), Lassosum (Lsum), and PRS-CS. PT computes the PRS on a subset of genetic variants based on LD-pruning and P-value thresholding. Lsum re-estimates the effect sizes using elastic net on GWAS summary statistics. The hyperparameters include the coefficients of L1 and L2 penalties. PRS-CS is a Bayesian polygenic prediction approach that uses a continuous shrinkage prior to derive posterior effect sizes. Overall, PT and Lsum are computationally fast while PRS-CS requires more computational time. In terms of prediction accuracy, Lsum and PRS-CS generally outperform PT ^5,6^.

### Transfer learning (TL-PRS) uses single ancestry GWAS summary statistics

Suppose that we have trained a PRS model using GWAS summary statistics from an ancestry group A, this model could be considered as prior knowledge to predict the genetic effects in the target ancestry group B. However, due to different LD patterns and possible effect size heterogeneity across ancestries, effect size estimation from ancestry group A can be viewed as biased estimators of effect sizes in ancestry group B. To adapt the model to the target population and achieve better prediction performance, we borrow the idea of transfer learning and attempt to combine information from the baseline model and the target sample data.

Specifically, for the target ancestry group, we have the following model:

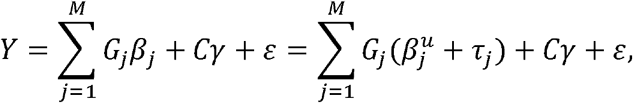

where *β*_*j*_ is the true effect size of the target ancestry group, assumed to be unknown; 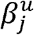 is given by the trained model; *τ*_*j*_ is the difference between 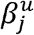 and *β*_*j*_; *C* is the covariate matrix including the intercept; and *γ* is a vector of covariate coefficients. Our goal is to minimize the following loss function:

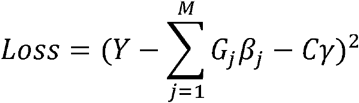

Since this problem can be converted into a convex optimization problem, we can perform a gradient descent algorithm on *β*_*j*_ with the initial value 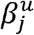. Given the current estimate *β*_*j*_^(*r*)^ in the *r*-th iteration, the next value, i.e. *β*_*j*_^(*r*+1)^ is

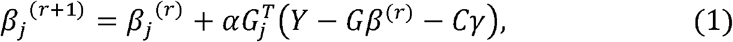

where *α* is the learning rate. The derivation can be found in Supplement 1. We note that 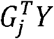 can be pre-calculated using the training data from the target ancestry group. In addition, early stopping of iteration is required to avoid overfitting.

Both the learning rate *α* and the number of iterations *n*_*stop*_ can be selected based on the validation dataset in terms of the best prediction accuracy. In order to reduce computation cost, we suggest choosing *α* from a small grid of values 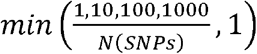, where *N* (*SNPs*) is the number of variants with non-zero effect sizes from the training model.

TL-PRS in Equation (1) requires individual level data for both model fitting and hyper-parameter tuning and we refer to it as TL-PRS (ind) specifically. When the individual level data from the training sample are not accessible, TL-PRS can still be applied if GWAS summary statistics of the target ancestry are available. In the step of model fitting, *G*^*T*^*Y* can be estimated by the summary statistics of the target ancestry^5^ and *G*^*T*^*G* can be estimated by the target ancestry using a public reference dataset, such as 1000 Genome Project. In the step of hyper-parameter tuning, the approach of using individual level data is applied in default. The model requirements of TL-PRS and TL-PRS(ind) can be found in Table S2.

### Combining multiple GWAS summary statistics from different ancestries

Suppose two PRSs, *PRS*_l_ and *PRS*_2_, are constructed from two different GWAS summary statistics, then the multi-ethnic PRS can be built as

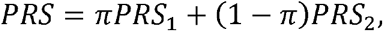

where *π* is a tuning parameter with range [0,1] and can be decided using the cross-validation method.^9^ This idea was first proposed by Márquez-Luna et al in 2017 using PT to construct a single-ancestry PRS, which was referred as PT-multi, and also used with PRS-CS (PRS-CSx^10^). This can be also used with Lsum, and we refer it Lsum-multi.

Similarly, the linear combination can also be applied to TL-PRS models. For example, with TL-PRS-Lsum models from two ancestries, we can linearly combine them first as the initial value and then implement transfer learning (referred as MTL-PRS-Lsum). MTL-PRS-CS can also be constructed in the same way.

Beyond the combination of two ancestries, we can further extend this idea to three or more different ancestries. Suppose that we have PRSs from three different ancestries *PRS*_l_, *PRS*_2_ and *PRS*_3_, then the multi-ethnic PRS can be built as

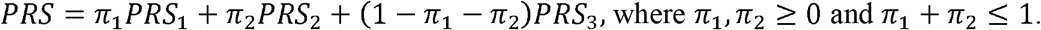

and then MTL-PRS can be constructed using linearly combined PRS as initial inputs.

### Simulations using South Asians samples in the UK Biobank

We simulated quantitative phenotypes using real 10,285 South Asian ancestry sample genotypes in UKBB. The proportion of causal markers was fixed as 0.1% and 1%, the SNP-heritability *h*_*g*_^2^ was fixed at 0.5. The normalized effect sizes *b*_*i*_ were generated from a normal distribution with mean 0 and variance equal to *h*_*g*_^2^ divided by the number of causal markers. The per-allele effect size is 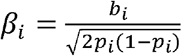, where *p*_*i*_ is the minor allele frequency of the *i*-th SNP. We simulated phenotypes as

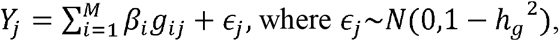

where *M* is the number of SNPs and only HapMap3 variants ^18^ were included in the simulation. The GWAS summary statistics based on 10,000 South Asian and 100,000 European ancestry individuals were generated respectively based on the formula 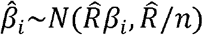, where n is the sample size and 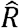 is the estimated correlation matrix of the LD block region using South Asian and European ancestry individuals from the 1000 Genomes Project. We assumed that causal variants could be shared across all ancestries (European and South Asian ancestries), but varying effect sizes were allowed and sampled from a multivariate normal distribution with a genetic correlation of 0.4, 0.7, or 1.0. Two sources (South Asian and European ancestry samples) of GWAS summary statistics were further generated and the sample sizes were 10,000 and 100,000, respectively. The simulation of the phenotype was repeated 20 times.

We randomly split the 10,285 simulated samples into training, validation, and testing datasets (Table S3). Ten PRS methods were included in our comparison, including single-source prediction methods (PT, Lsum, TL-PRS-Lsum, PRS-CS, and TL-PRS-CS) and multi-source prediction methods (PT-multi, Lsum-multi, MTL-PRS-Lsum, PRS-CSx, and MTL-PRS-CS). Their predictive performances were measured by *R*^2^ between the simulated and predicted phenotypes in the testing dataset.

Although TL-PRS doesn’t require individual-level data of the training dataset, TL-PRS requires individual level validation data. For a fair comparison, we applied the PRS baseline models (PT, Lsum, PRS-CS) using the combination of training and validation datasets for validation. Among them, PT and Lsum requires individual level data while PRS-CS does not. PT-multi, Lsum-multi and PRS-CSx were then implemented by linearly combining PT, Lsum, and PRS-CS models, respectively. We note that when selecting the tuning parameter *π*, PRS-CSx also requires individual level data. For the TL-PRS methods, SAIGE^19^ was used first on the training dataset to calculate GWAS summary statistics, and baseline models (Lsum, Lsum-multi, PRS-CS and PRS-CSx) were pre-trained using only training dataset. Based on the calculated summary statistics and pre-trained models, TL-PRS can further be fine-tuned given the individual level data of the validation dataset. We note that TL-PRS doesn’t require individual level data for training. The implementation details of all methods can be found in Table S1.

### Analysis of South Asian and African ancestry samples in the UK Biobank

We constructed PRSs for following target samples in UK Biobank: South Asian (SAS), and African (AFR) ancestry individuals. In each target sample, we used the software KING to exclude one individual in each related pair up to second-degree relatives. We then built the polygenic prediction models on the following eight traits: high-density lipoproteins (HDL), low-density lipoproteins (LDL), body mass index (BMI), triglycerides (TG), systolic blood pressure (SBP), diastolic blood pressure (DBP), coronary artery disease (CAD), and Type 2 diabetes (T2D). The first six traits were quantitative and the last two traits were dichotomous.

Summary statistics of GWAS analyses on White British in UK Biobank (UKBB) and Japanese in Biobank Japan (BBJ) were downloaded from UKBB (https://pheweb.org/UKB-Neale/) and BBJ PheWeb (http://jenger.riken.jp/en/result). We restricted our analysis to common variants (MAF>=0.01) presented in summary data and target genotype files after removing A/T and C/G SNPs to eliminate potential strand ambiguity^9^.

For each ancestry, the target samples were randomly split into a training dataset, a validation dataset, and a testing dataset (Table S3). We followed the same strategy of training models as the simulation (Table S1). We applied single-source prediction methods (PT, Lsum, TL-PRS-Lsum, PRS-CS, TL-PRS-CS) to UKBB and BBJ summary statistics and used multi-source prediction methods (PT-multi, Lsum-multi, MTL-PRS-Lsum, PRS-CSx, MTL-PRS-CS) to combine UKBB and BBJ GWAS results. The prediction accuracy was assessed in the testing dataset of each target ancestry separately, adjusting for age, sex and the top four principal components (PCs). We used *R*^2^ as the prediction accuracy metric for quantitative traits and Nagelkerke *R*^2^ for dichotomous traits.

## Data Availability

All data produced are available online at https://www.ukbiobank.ac.uk/.

http://jenger.riken.jp/en/result

https://www.internationalgenome.org/

## ACKNOWLEDGEMENTS

This research has been conducted using the UK Biobank Resource under application number 45227. We thank Biobank Japan (BBJ) for releasing the genome-wide association summary statistics. B.M is supported by NSF DMS1712933 and NIH R01HG008773. S.L.is supported by BP+ Program through NRF of Korea funded by the Ministry of Science and ICT (2020H1D3A2A03100666). Z.Z. is supported by NIH R01HG008773 and R01LM012535.

## WEB RESOURCES

UK Biobank: https://www.ukbiobank.ac.uk/

BBJ summary statistics: http://jenger.riken.jp/en/result

1000 Genome Project: https://www.internationalgenome.org/

KING software: https://www.kingrelatedness.com/manual.shtml

Lassosum: https://github.com/tshmak/lassosum

PRS-CS: https://github.com/getian107/PRScs

PRS-CSx: https://github.com/getian107/PRScsx

TL-PRS: https://github.com/ZhangchenZhao/TLPRS

