## Supplementary Material for "The Construction of Multi-ethnic Polygenic Risk Score using Transfer Learning"

### Supplementary Materials

**Supplement 1.** Derivation of gradient descent algorithm (Equation 1)

When carrying out gradient descent algorithm, our goal is to minimize the following loss function:

$$Loss = (Y - G\beta - C\gamma)^2.$$

with respect to $\beta_j$ for each $j$ in turn. The first and second derivative of loss function is

$$\frac{\partial Loss}{\partial \beta_j} = -2G_j^T(Y - G\beta - C\gamma), \qquad \frac{\partial^2 Loss}{\partial \beta_j^2} = 2G_j^T G > 0.$$

Since the second derivative is constantly larger than 0, the iterative formula of gradient descent can be used:

$$\beta_j{}^{(r+1)} = \beta_j{}^{(r)} - \alpha' \frac{\partial Loss}{\partial \beta_j} = \beta_j{}^{(r)} + 2\alpha' G_j^T\left(Y - G\beta^{(i)} - C\gamma\right),$$

and we define the learning rate $\alpha = 2\alpha'$.

### Supplementary Figures

**Figure S1.** Prediction accuracy of TL-PRS(ind) and TL-PRS methods in simulations. Different genetic designs (0.1% and 1% causal variants) were simulated as well as cross-population genetic correlations (0.4, 0.7 and 1.0). Heritability was fixed at 50%. Prediction accuracy was measured by the squared correlation (R2) between the simulated and predicted phenotypes in the testing dataset, averaged across 20 simulation replicates. Error bar indicates the standard deviation of R2 across simulation replicates.

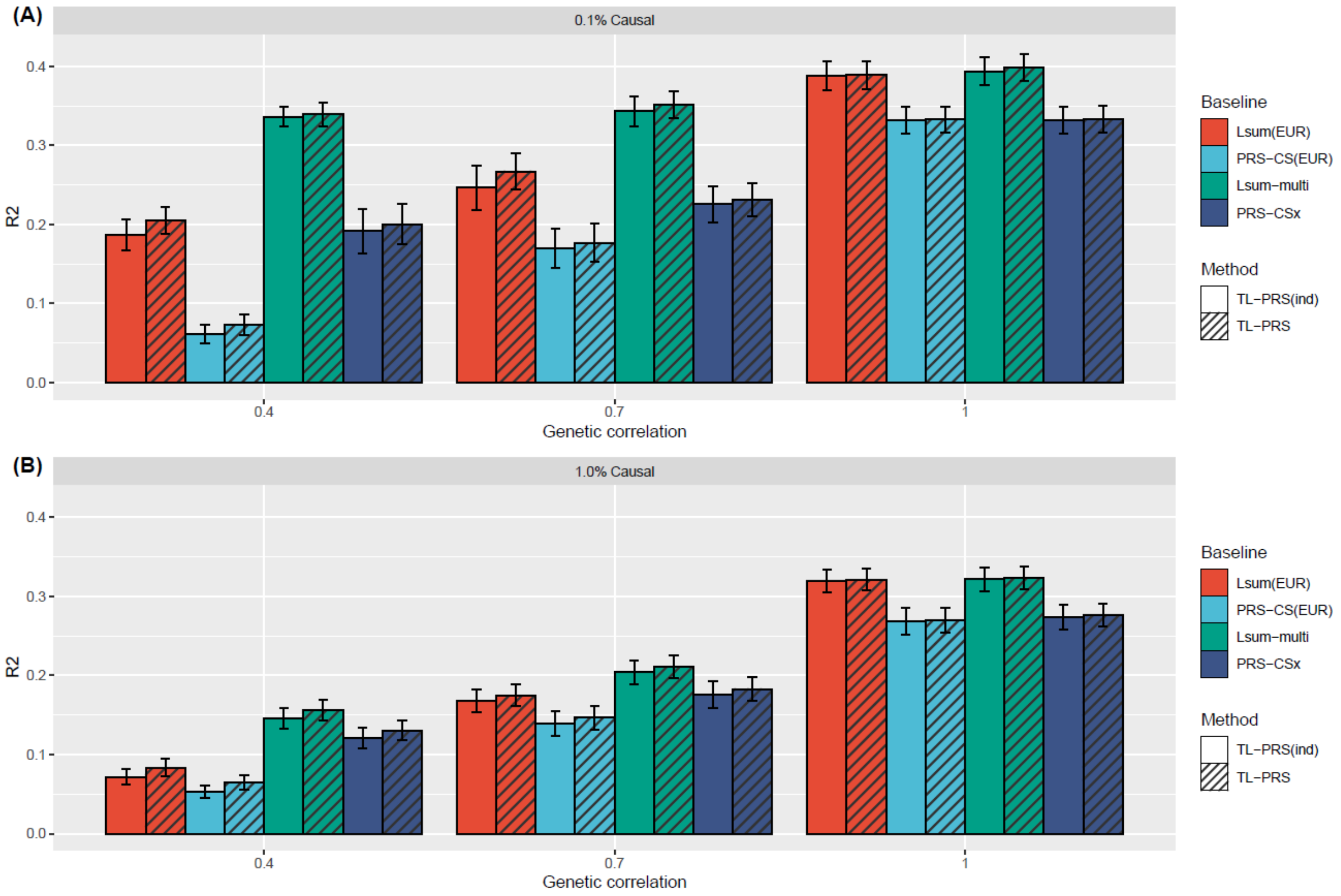

**Figure S2.** Cumulant event plot in terms of the top 10% PRS constructed by transfer learning methods and their baseline methods.

(a) South Asian, Type 2 diabetes, Case: Control=419:2211

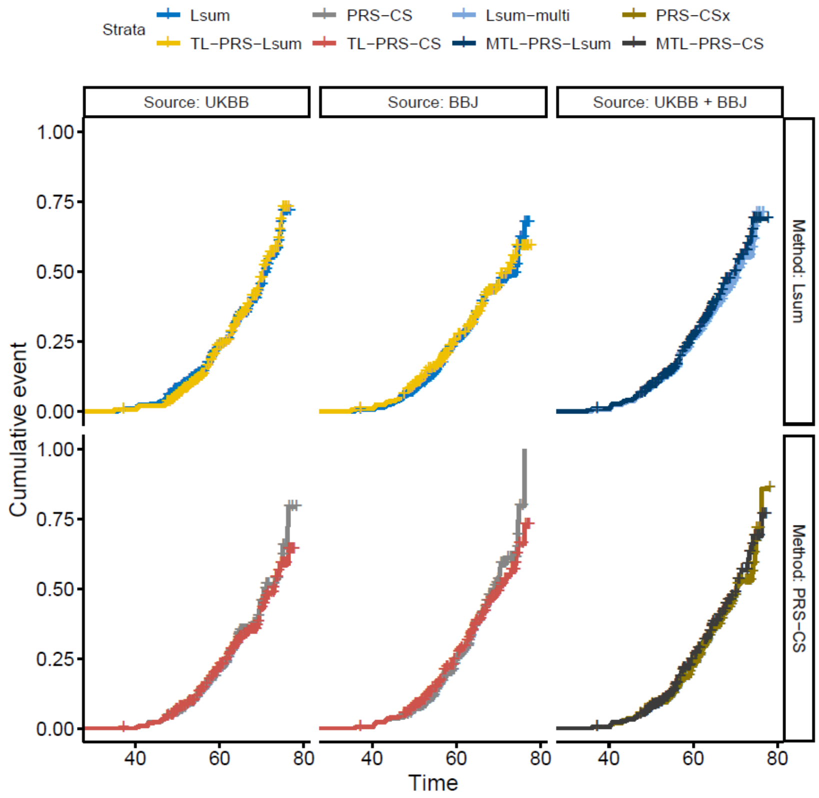


(b) South Asian, Coronary artery disease, Case: Control=362:2270

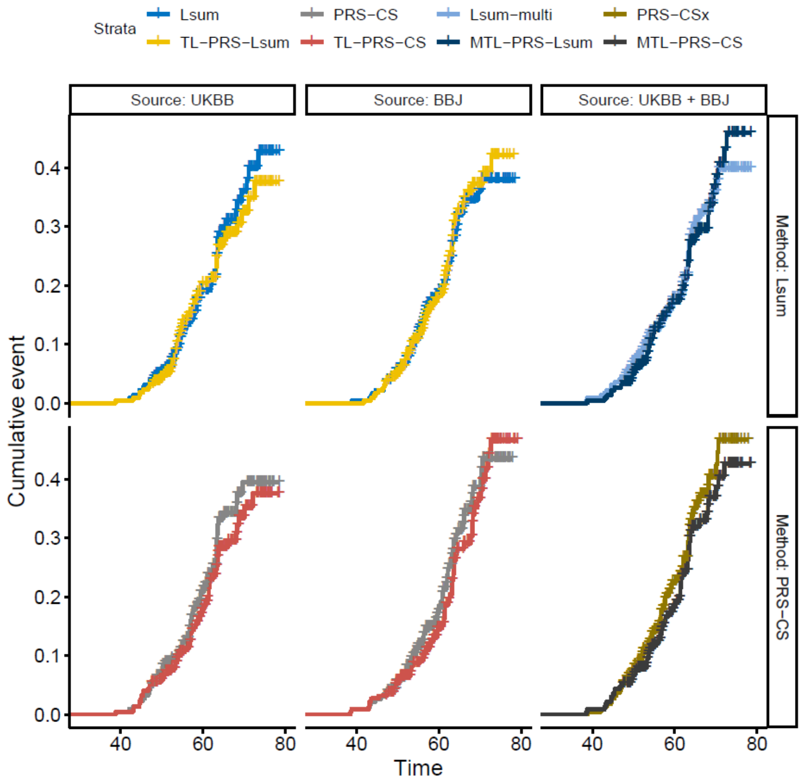

(c) African, Type 2 diabetes, Case: Control=177:1812

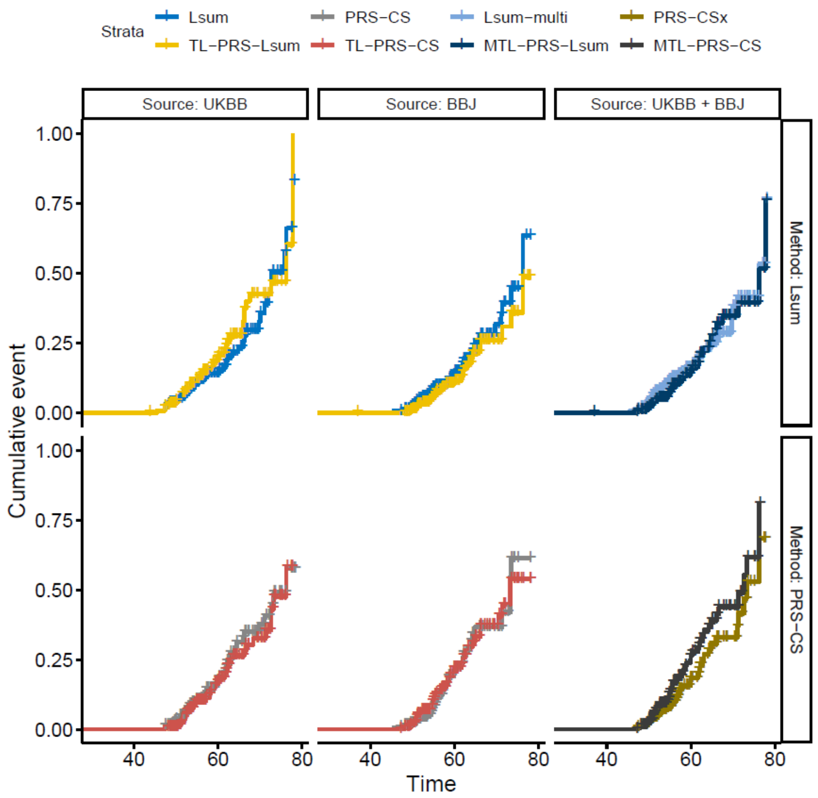


(d) African, Coronary artery disease, Case: Control=94:1902

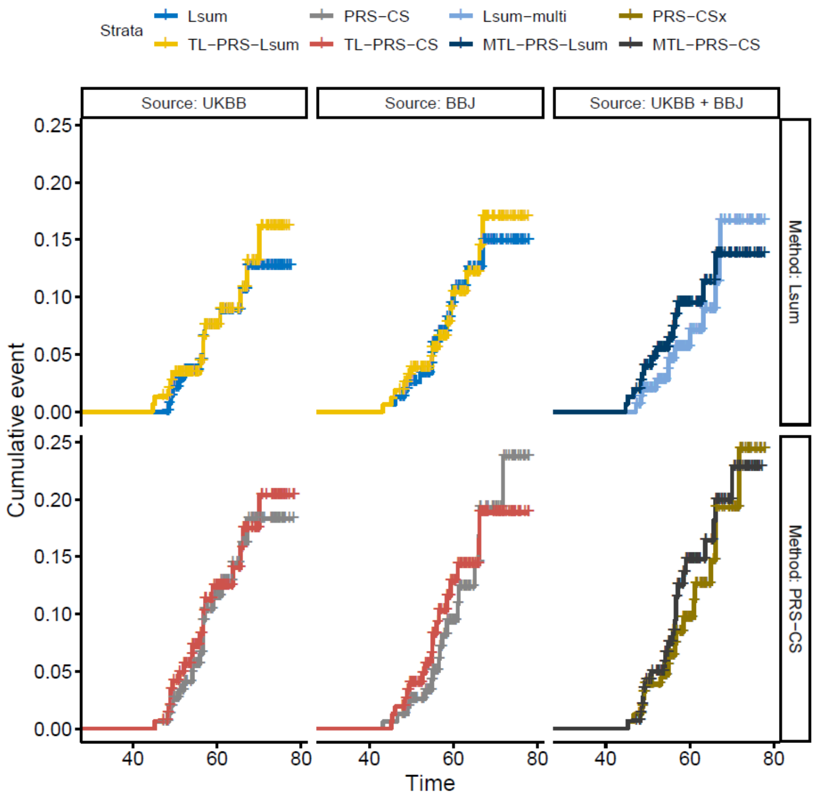

### Supplementary Tables

**Table S1. The implementation of prediction methods in the simulation and application of UK Biobank**

**(a)** The implementation of single-source prediction methods.

| Method | Training data | Validation data | Testing data |
|---|---|---|---|
| PT | Select the hyperparameters using the combination of training and validation data. Individual-level data are required. | | Assess prediction performance using individual-level data |
| Lsum | Select the hyperparameters using the combination of training and validation data. Individual-level data are required. | | Assess prediction performance using individual-level data |
| PRS-CS | Select the hyperparameters using the combination of training and validation data. Individual-level data are **not** required. | | Assess prediction performance using individual-level data |
| TL-PRS-Lsum | Train the baseline Lsum model using training data, and then implement TL-PRS-Lsum. Individual-level data are **not** required. | Fine-tune TL-PRS-Lsum model using validation data.<br>Individual-level data are recommended. | Assess prediction performance using individual-level data |
| TL-PRS-CS | Train the baseline PRS-CS model using training data, and then implement TL-PRS-CS. Individual-level data are **not** required. | Fine-tune TL-PRS-CS model using validation data.<br>Individual-level data are recommended. | Assess prediction performance using individual-level data |

**(b)** The implementation of multi-source prediction methods.

| Method | Training data | Validation data | Testing data |
|---|---|---|---|
| PT-multi<br>Lsum-multi<br>PRS-CSx | Select the weights (hyperparameter) to linearly combine single-source prediction models using combination of training and validation data.<br>Individual-level data are required to fine-tune the weight parameter. | | Assess prediction performance using individual-level data |
| MTL-PRS-Lsum<br>MTL-PRS-CS | Train the baseline Lsum-multi/PRS-CSx model using training data, and then implement TL-PRS. Individual-level data are **not** required. | Fine-tune TL-PRS model using validation data.<br>Individual-level data are recommended. | Assess prediction performance using individual-level data |

**Table S2.** List of data sets used in simulations and analyses of real phenotypes.

| Target Population | Trait | Total sample size | Training sample size | Validation sample size | Testing sample size |
|---|---|---|---|---|---|
| South Asian (SAS) | Simulation and real phenotypes | 10,285 | 5,000 | 2,635 | 2,635 |
| African (AFR) | real phenotypes | 8,168 | 4,000 | 2,169 | 1,999 |

**Table S3.** The model requirement of TL-PRS and TL-PRS (ind).

| Methods | Training dataset | Validation dataset | Testing dataset |
|---|---|---|---|
| TL-PRS | Only require summary statistics | Individual-level data are recommended. | Require individual-level data to assess prediction performance |
| TL-PRS (ind) | Require individual-level data | Require individual-level data | Require individual-level data to assess prediction performance |

**Table S4.** Prediction accuracy of different PRS construction methods in analyses of eight traits in the South Asian cohort of UK Biobank. The approach with highest prediction $R^2$ is highlighted with bold.

(a) HDL

| Model | Prediction R2 of PRS | Mean difference between top 10% and bottom 10% PRS |
|---|---|---|
| PT (UKBB) | 0.07 | 0.301 |
| Lsum (UKBB) | 0.092 | 0.323 |
| TL-PRS-Lsum (UKBB) | 0.099 | 0.355 |
| PRS-CS (UKBB) | 0.101 | 0.332 |
| TL-PRS-CS (UKBB) | 0.109 | 0.376 |
| PT (BBJ) | 0.044 | 0.225 |
| Lsum (BBJ) | 0.061 | 0.291 |
| TL-Lsum (BBJ) | 0.065 | 0.293 |
| PRS-CS (BBJ) | 0.046 | 0.208 |
| TL-PRS-CS (BBJ) | 0.062 | 0.276 |
| PT-multi | 0.082 | 0.328 |
| Lsum-multi | 0.105 | 0.367 |
| MTL-PRS-Lsum | 0.111 | 0.38 |
| PRSCSx | 0.107 | 0.344 |
| **MTL-PRS-CS** | **0.117** | **0.369** |

(b) LDL

| Model | Prediction R2 of PRS | Mean difference between top 10% and bottom 10% PRS |
|---|---|---|
| PT (UKBB) | 0.02 | 0.448 |
| Lsum (UKBB) | 0.023 | 0.534 |
| TL-PRS-Lsum (UKBB) | 0.024 | 0.575 |
| PRS-CS (UKBB) | 0.02 | 0.429 |
| TL-PRS-CS (UKBB) | 0.024 | 0.509 |
| PT (BBJ) | 0.012 | 0.243 |
| Lsum (BBJ) | 0.019 | 0.432 |
| TL-Lsum (BBJ) | 0.019 | 0.48 |
| PRS-CS (BBJ) | 0.007 | 0.22 |
| TL-PRS-CS (BBJ) | 0.013 | 0.383 |
| PT-multi | 0.024 | 0.548 |
| Lsum-multi | 0.03 | 0.543 |
| **MTL-PRS-Lsum** | **0.031** | **0.533** |
| PRSCSx | 0.022 | 0.475 |
| MTL-PRS-CS | 0.026 | 0.425 |

(c) BMI

| Model | Prediction R2 of PRS | Mean difference between top 10% and bottom 10% PRS |
|---|---|---|
| PT (UKBB) | 0.047 | 3.025 |
| Lsum (UKBB) | 0.072 | 3.918 |
| TL-PRS-Lsum (UKBB) | 0.073 | 4.053 |
| PRS-CS (UKBB) | 0.067 | 3.972 |
| TL-PRS-CS (UKBB) | 0.073 | 4.155 |
| PT (BBJ) | 0.017 | 1.951 |
| Lsum (BBJ) | 0.022 | 2.404 |
| TL-Lsum (BBJ) | 0.025 | 2.633 |
| PRS-CS (BBJ) | 0.023 | 2.083 |
| TL-PRS-CS (BBJ) | 0.028 | 2.072 |
| PT-multi | 0.055 | 3.754 |
| **Lsum-multi** | **0.077** | **4.157** |
| MTL-PRS-Lsum | 0.076 | 4.162 |
| PRSCSx | 0.071 | 3.817 |
| MTL-PRS-CS | 0.075 | 4.291 |

(d) TG

| Model | Prediction R2 of PRS | Mean difference between top 10% and bottom 10% PRS |
|---|---|---|
| PT (UKBB) | 0.054 | 0.899 |
| Lsum (UKBB) | 0.082 | 1.142 |
| TL-PRS-Lsum (UKBB) | 0.081 | 1.119 |
| PRS-CS (UKBB) | 0.083 | 1.047 |
| TL-PRS-CS (UKBB) | 0.082 | 1.016 |
| PT (BBJ) | 0.046 | 0.77 |
| Lsum (BBJ) | 0.065 | 0.949 |
| TL-Lsum (BBJ) | 0.069 | 0.95 |
| PRS-CS (BBJ) | 0.053 | 0.896 |
| TL-PRS-CS (BBJ) | 0.053 | 0.856 |
| PT-multi | 0.072 | 0.958 |
| **Lsum-multi** | **0.094** | **1.162** |
| MTL-PRS-Lsum | 0.088 | 1.143 |
| PRSCSx | 0.093 | 1.059 |
| MTL-PRS-CS | 0.09 | 1.069 |

(e) SBP

| Model | Prediction R2 of PRS | Mean difference between top 10% and bottom 10% PRS |
|---|---|---|
| PT (UKBB) | 0.021 | 10.788 |
| Lsum (UKBB) | 0.035 | 13.902 |
| TL-PRS-Lsum (UKBB) | 0.038 | 16.129 |
| PRS-CS (UKBB) | 0.036 | 12.569 |
| TL-PRS-CS (UKBB) | 0.035 | 15.075 |
| PT (BBJ) | 0.005 | 6.824 |
| Lsum (BBJ) | 0.01 | 8.694 |
| TL-Lsum (BBJ) | 0.011 | 7.373 |
| PRS-CS (BBJ) | 0.008 | 7.161 |
| TL-PRS-CS (BBJ) | 0.009 | 6.145 |
| PT-multi | 0.023 | 12.404 |
| Lsum-multi | 0.038 | 14.231 |
| **MTL-PRS-Lsum** | **0.042** | **16.455** |
| PRSCSx | 0.038 | 14.306 |
| MTL-PRS-CS | 0.039 | 15.322 |

(f) DBP

| Model | Prediction R2 of PRS | Mean difference between top 10% and bottom 10% PRS |
|---|---|---|
| PT (UKBB) | 0.013 | 4.376 |
| Lsum (UKBB) | 0.034 | 6.667 |
| TL-PRS-Lsum (UKBB) | 0.034 | 6.914 |
| PRS-CS (UKBB) | 0.035 | 5.91 |
| TL-PRS-CS (UKBB) | 0.036 | 7.098 |
| PT (BBJ) | 0.001 | 0.973 |
| Lsum (BBJ) | 0.003 | 1.855 |
| TL-Lsum (BBJ) | 0.004 | 2.012 |
| PRS-CS (BBJ) | 0.003 | 3.145 |
| TL-PRS-CS (BBJ) | 0.006 | 2.498 |
| PT-multi | 0.013 | 3.396 |
| Lsum-multi | 0.033 | 6.227 |
| MTL-PRS-Lsum | 0.035 | 7.529 |
| PRSCSx | 0.034 | 6.404 |
| **MTL-PRS-CS** | **0.036** | **7.573** |

(g) CAD

| Model | Prediction pseudo R2 of PRS | AUC | Risk ratio between top 10% and bottom 10% PRS |
|---|---|---|---|
| PT (UKBB) | 0.014 | 0.747 | 2.381 |
| Lsum (UKBB) | 0.023 | 0.752 | 2.227 |
| TL-PRS-Lsum (UKBB) | 0.021 | 0.751 | 2.25 |
| PRS-CS (UKBB) | 0.022 | 0.752 | 4.231 |
| TL-PRS-CS (UKBB) | 0.031 | 0.756 | 3 |
| PT (BBJ) | 0.007 | 0.743 | 1.68 |
| Lsum (BBJ) | 0.022 | 0.75 | 2.611 |
| TL-Lsum (BBJ) | 0.018 | 0.75 | 2.273 |
| PRS-CS (BBJ) | 0.023 | 0.751 | 2.8 |
| TL-PRS-CS (BBJ) | 0.032 | 0.757 | 3.067 |
| PT-multi | 0.019 | 0.751 | 2.882 |
| Lsum-multi | 0.036 | 0.758 | 2.526 |
| MTL-PRS-Lsum | 0.029 | 0.756 | 3.267 |
| PRSCSx | 0.036 | 0.758 | 3.688 |
| **MTL-PRS-CS** | **0.043** | **0.763** | **5** |

(h) Type II diabetes

| Model | Prediction pseudo R2 of PRS | AUC | Risk ratio between top 10% and bottom 10% PRS |
|---|---|---|---|
| PT (UKBB) | 0.012 | 0.692 | 2.44 |
| Lsum (UKBB) | 0.031 | 0.708 | 2.8 |
| TL-PRS-Lsum (UKBB) | 0.033 | 0.708 | 2.917 |
| PRS-CS (UKBB) | 0.033 | 0.71 | 2.68 |
| TL-PRS-CS (UKBB) | 0.037 | 0.712 | 3.238 |
| PT (BBJ) | 0.014 | 0.693 | 1.867 |
| Lsum (BBJ) | 0.03 | 0.707 | 2.75 |
| TL-Lsum (BBJ) | 0.029 | 0.707 | 4.062 |
| PRS-CS (BBJ) | 0.035 | 0.708 | 2.846 |
| TL-PRS-CS (BBJ) | 0.047 | 0.717 | 3.889 |
| PT-multi | 0.023 | 0.701 | 2.583 |
| Lsum-multi | 0.048 | 0.72 | 4.688 |
| MTL-PRS-Lsum | 0.049 | 0.72 | 8.444 |
| PRSCSx | 0.048 | 0.719 | 3.217 |
| **MTL-PRS-CS** | **0.056** | **0.723** | **5.286** |

**Table S5.** Prediction accuracy of different PRS construction methods in analyses of eight traits in the African cohort of UK Biobank. The approach with highest prediction $R^2$ is highlighted with bold.

(a) HDL

| Model | Prediction R2 of PRS | Mean difference between top 10% and bottom 10% PRS |
|---|---|---|
| PT (UKBB) | 0.045 | 0.228 |
| Lsum (UKBB) | 0.042 | 0.287 |
| TL-PRS-Lsum (UKBB) | 0.045 | 0.31 |
| PRS-CS (UKBB) | 0.05 | 0.236 |
| TL-PRS-CS (UKBB) | 0.054 | 0.265 |
| PT (BBJ) | 0.035 | 0.259 |
| Lsum (BBJ) | 0.047 | 0.345 |
| TL-Lsum (BBJ) | 0.05 | 0.347 |
| PRS-CS (BBJ) | 0.047 | 0.265 |
| TL-PRS-CS (BBJ) | 0.048 | 0.325 |
| PT-multi | 0.052 | 0.24 |
| Lsum-multi | 0.062 | 0.341 |
| MTL-PRS-Lsum | 0.064 | 0.317 |
| PRSCSx | 0.071 | 0.321 |
| **MTL-PRS-CS** | **0.072** | **0.33** |

(b) LDL

| Model | Prediction R2 of PRS | Mean difference between top 10% and bottom 10% PRS |
| --- | --- | --- |
| PT (UKBB) | 0.012 | 0.388 |
| Lsum (UKBB) | 0.033 | 0.625 |
| TL-PRS-Lsum (UKBB) | 0.058 | 0.779 |
| PRS-CS (UKBB) | 0.022 | 0.552 |
| TL-PRS-CS (UKBB) | 0.028 | 0.506 |
| PT (BBJ) | 0.028 | 0.474 |
| Lsum (BBJ) | 0.048 | 0.595 |
| TL-Lsum (BBJ) | 0.068 | 0.795 |
| PRS-CS (BBJ) | 0.023 | 0.421 |
| TL-PRS-CS (BBJ) | 0.028 | 0.565 |
| PT-multi | 0.03 | 0.531 |
| Lsum-multi | 0.052 | 0.727 |
| **MTL-PRS-Lsum** | **0.068** | **0.926** |
| PRSCSx | 0.037 | 0.662 |
| MTL-PRS-CS | 0.044 | 0.721 |

(c) BMI

| Model | Prediction R2 of PRS | Mean difference between top 10% and bottom 10% PRS |
|---|---|---|
| PT (UKBB) | 0.028 | 3.416 |
| Lsum (UKBB) | 0.038 | 3.762 |
| TL-PRS-Lsum (UKBB) | 0.039 | 4.035 |
| PRS-CS (UKBB) | 0.032 | 3.48 |
| TL-PRS-CS (UKBB) | 0.034 | 3.399 |
| PT (BBJ) | 0.008 | 1.242 |
| Lsum (BBJ) | 0.013 | 1.911 |
| TL-Lsum (BBJ) | 0.013 | 2.466 |
| PRS-CS (BBJ) | 0.012 | 1.946 |
| TL-PRS-CS (BBJ) | 0.02 | 2.677 |
| PT-multi | 0.031 | 3 |
| **Lsum-multi** | **0.043** | **4.018** |
| MTL-PRS-Lsum | 0.043 | 3.676 |
| PRSCSx | 0.036 | 4.006 |
| MTL-PRS-CS | 0.042 | 4.057 |

(d) TG

| Model | Prediction R2 of PRS | Mean difference between top 10% and bottom 10% PRS |
|---|---|---|
| PT (UKBB) | 0.006 | 0.172 |
| Lsum (UKBB) | 0.011 | 0.271 |
| TL-PRS-Lsum (UKBB) | 0.011 | 0.263 |
| PRS-CS (UKBB) | 0.015 | 0.245 |
| TL-PRS-CS (UKBB) | 0.019 | 0.409 |
| PT (BBJ) | 0.006 | 0.207 |
| Lsum (BBJ) | 0.009 | 0.167 |
| TL-Lsum (BBJ) | 0.009 | 0.177 |
| PRS-CS (BBJ) | 0.004 | 0.138 |
| TL-PRS-CS (BBJ) | 0.008 | 0.209 |
| PT-multi | 0.009 | 0.233 |
| Lsum-multi | 0.013 | 0.267 |
| MTL-PRS-Lsum | 0.011 | 0.21 |
| PRSCSx | 0.016 | 0.257 |
| **MTL-PRS-CS** | **0.02** | **0.378** |

(e) SBP

| Model | Prediction R2 of PRS | Mean difference between top 10% and bottom 10% PRS |
|---|---|---|
| PT (UKBB) | 0.003 | 4.451 |
| Lsum (UKBB) | 0.011 | 7.015 |
| TL-PRS-Lsum (UKBB) | 0.011 | 7.015 |
| PRS-CS (UKBB) | 0.012 | 8.769 |
| **TL-PRS-CS (UKBB)** | **0.013** | **9.677** |
| PT (BBJ) | 0.001 | 3.39 |
| Lsum (BBJ) | 0.001 | 3.215 |
| TL-Lsum (BBJ) | 0.002 | 1.544 |
| PRS-CS (BBJ) | 0.004 | 4.379 |
| TL-PRS-CS (BBJ) | 0.003 | 2.579 |
| PT-multi | 0.003 | 4.826 |
| Lsum-multi | 0.011 | 7.113 |
| MTL-PRS-Lsum | 0.011 | 6.713 |
| PRSCSx | 0.013 | 8.61 |
| MTL-PRS-CS | 0.012 | 7.897 |

(f) DBP

| Model | Prediction R2 of PRS | Mean difference between top 10% and bottom 10% PRS |
|---|---|---|
| PT (UKBB) | 0.001 | 1.872 |
| Lsum (UKBB) | 0.004 | 2.579 |
| TL-PRS-Lsum (UKBB) | 0.004 | 2.579 |
| PRS-CS (UKBB) | 0.006 | 2.456 |
| **TL-PRS-CS (UKBB)** | **0.009** | **3.338** |
| PT (BBJ) | 0 | 1.046 |
| Lsum (BBJ) | 0.003 | 1.913 |
| TL-Lsum (BBJ) | 0.003 | 1.785 |
| PRS-CS (BBJ) | 0.002 | 2.082 |
| TL-PRS-CS (BBJ) | 0.002 | 3.39 |
| PT-multi | 0.002 | 2.523 |
| Lsum-multi | 0.006 | 2.426 |
| MTL-PRS-Lsum | 0.006 | 2.713 |
| PRSCSx | 0.007 | 3.944 |
| MTL-PRS-CS | 0.009 | 3.19 |

(g) CAD

| Model | Prediction pseudo R2 of PRS | AUC of | Risk ratio between top 10% and bottom 10% PRS |
|---|---|---|---|
| **PT (UKBB)** | **0.028** | **0.72** | **3.167** |
| Lsum (UKBB) | 0.003 | 0.694 | 4 |
| TL-PRS-Lsum (UKBB) | 0.01 | 0.704 | 2.6 |
| PRS-CS (UKBB) | 0.01 | 0.705 | 4.25 |
| TL-PRS-CS (UKBB) | 0.016 | 0.71 | 3.6 |
| PT (BBJ) | 0.002 | 0.693 | 1.556 |
| Lsum (BBJ) | 0.003 | 0.695 | 3.5 |
| TL-Lsum (BBJ) | 0.004 | 0.695 | 3 |
| PRS-CS (BBJ) | 0.006 | 0.698 | 1.75 |
| TL-PRS-CS (BBJ) | 0.005 | 0.696 | 2.5 |
| PT-multi | 0.019 | 0.713 | 4.667 |
| Lsum-multi | 0.005 | 0.697 | 2.4 |
| MTL-PRS-Lsum | 0.008 | 0.699 | 2.167 |
| PRSCSx | 0.009 | 0.701 | 2 |
| MTL-PRS-CS | 0.02 | 0.712 | 5 |

(h) Type II diabetes

| Model | Prediction pseudo R2 of PRS | AUC | Risk ratio between top 10% and bottom 10% PRS |
|---|---|---|---|
| PT (UKBB) | 0.005 | 0.734 | 1.6 |
| Lsum (UKBB) | 0.012 | 0.74 | 1.5 |
| TL-PRS-Lsum (UKBB) | 0.009 | 0.741 | 1.706 |
| PRS-CS (UKBB) | 0.022 | 0.745 | 2.455 |
| TL-PRS-CS (UKBB) | 0.022 | 0.748 | 4.5 |
| PT (BBJ) | 0.005 | 0.735 | 1.643 |
| Lsum (BBJ) | 0.003 | 0.732 | 1.533 |
| TL-Lsum (BBJ) | 0.002 | 0.732 | 1.636 |
| PRS-CS (BBJ) | 0.007 | 0.732 | 1.923 |
| TL-PRS-CS (BBJ) | 0.018 | 0.739 | 2.357 |
| PT-multi | 0.009 | 0.737 | 1.412 |
| Lsum-multi | 0.011 | 0.738 | 1.643 |
| MTL-PRS-Lsum | 0.012 | 0.739 | 1.643 |
| PRSCSx | 0.021 | 0.74 | 1.6 |
| **MTL-PRS-CS** | **0.026** | **0.747** | **3.5** |

Table S6. The top three methods for all 8 traits in the South Asian and African ancestries in terms of prediction $R^2$. Single-source prediction methods (PT, Lsum, TL-PRS-Lsum, PRS-CS, TL-PRS-CS) based on UKBB and BBJ GWAS results and mulit-source PRS methods (PT-multi, Lsum-multi, MTL-PRS-Lsum, PRS-CSx, MTL-PRS-CS) were included in the comparison and our approaches were highlighted using bold.

| Target population | trait | Best approach (rank 1) | Rank 2 | Rank 3 |
|---|---|---|---|---|
| **South Asian** | HDL | **MTL-PRS-CS** | **MTL-PRS-Lsum** | **TL-PRS-CS(UKBB)** |
| | LDL | **MTL-PRS-Lsum** | Lsum-multi | **MTL-PRS-CS** |
| | BMI | Lsum-multi | **MTL-PRS-Lsum** | **MTL-PRS-CS** |
| | TG | Lsum-multi | PRS-CSx | **MTL-PRS-CS** |
| | SBP | **MTL-PRS-Lsum** | **MTL-PRS-CS** | PRS-CSx |
| | DBP | **MTL-PRS-CS** | **TL-PRS-CS(UKBB)** | **MTL-PRS-Lsum** |
| | CAD | **MTL-PRS-CS** | PRS-CSx | Lsum-multi |
| | T2D | **MTL-PRS-CS** | **MTL-PRS-Lsum** | PRS-CSx |
| **African** | HDL | **MTL-PRS-CS** | PRS-CSx | **MTL-PRS-Lsum** |
| | LDL | **TL-PRS-Lsum(BBJ)** | **MTL-PRS-Lsum** | **TL-PRS-Lsum(UKBB)** |
| | BMI | **MTL-PRS-Lsum** | Lsum-multi | **MTL-PRS-CS** |
| | TG | **MTL-PRS-CS** | **TL-PRS-CS(UKBB)** | PRS-CSx |
| | SBP | **TL-PRS-CS(UKBB)** | PRS-CSx | **MTL-PRS-CS** |
| | DBP | **MTL-PRS-CS** | **TL-PRS-CS(UKBB)** | PRS-CSx |
| | CAD | PT(UKBB) | **MTL-PRS-CS** | PT-multi |
| | T2D | **MTL-PRS-CS** | PRS-CS(UKBB) | **TL-PRS-CS(UKBB)** |
